# SARS-CoV-2 Spike Protein Receptor Binding-ACE2 Interaction Increases Carbohydrate Sulfotransferases and Reduces N-Acetylgalactosamine-4-Sulfatase through Phospho-p38-MAPK and RB-E2F1

**DOI:** 10.1101/2023.01.24.23284890

**Authors:** Sumit Bhattacharyya, Joanne K. Tobacman

**Affiliations:** Jesse Brown VA Medical Center and University of Illinois at Chicago, Chicago, IL 60612 U.S.A.

## Abstract

Immunohistochemistry of post-mortem lung tissue from patients with SARS-CoV2 infection showed marked decline in intensity and distribution of N-acetylgalactosamine-4-sulfatase (Arylsulfatase B; ARSB), increase of total chondroitin sulfate by immunohistochemistry, and increase of vascular-associated carbohydrate sulfotransferase (CHST)15 [1]. The mechanisms leading to these observations were not explained by signaling pathways known to be activated by exposure to coronaviruses. This report addresses the underlying reactions leading to these observations in a cell-based model, using normal, human, primary small airway epithelial cells, treated with the SARS-CoV-2 spike protein receptor binding domain protein.

## Introduction

Chondroitin sulfates are vital components of cells and the extracellular matrix (ECM) in all human tissues and throughout living organisms. Biosynthesis of CSE by CHST15 proceeds by transfer of the sulfate residue from 3′-phosphoadenosine-5′-phosphosulfate (PAPS) to the 6-OH of GalNAc4S of C4S [2]. Mechanisms for enhanced CHST15 or CHST11 expression or for decline in ARSB following viral infection have not been reported previously, although the role of sulfated glycosaminoglycans, particularly heparin and heparan sulfate, in SARS-CoV-2 viral uptake and in inhibition of viral infection of human cells has been investigated [3–5].

CHST15 [N-acetylgalactosamine 4-sulfate 6-O-Sulfotransferase; B-cell RAG (Recombination Activating Gene)-associated protein; GALNAc4S-6ST] is required for the synthesis of chondroitin sulfate E (CSE) which is composed of alternating β-1,4 and β-1,3 linked D-glucuronate and D-N-acetylgalactosamine-4,6-sulfate residues. Increases in CHST15 or in CSE, have been associated with increased fibrosis in cardiac, lung, and other tissues and with infectivity of dengue virus [6–11], as well as in several malignant cells and tissues, including pancreas, ovary, colon, lung, and brain [12–16]. CHST11 is a carbohydrate sulfotransferase that adds 4-sulfate groups to D-N-acetylgalactosamine residues which are linked to D-glucuronate by β-1,3 glycosidic bonds in chondroitin 4-sulfate or to D-iduronate in dermatan sulfate. Increases in chondroitin 4-sulfate (C4S) and dermatan sulfate follow decline in arylsulfatase B (ARSB, N-acetylgalactosamine-4-sulfatase), since ARSB is the enzyme that removes 4-sulfate groups from N-acetylgalactosamine 4-sulfate residues and is required for the degradation of C4S and dermatan sulfate, as evident by the accumulation of these sulfated glycosaminoglycans in the congenital disease Mucopolysaccharidosis VI (Maroteaux-Lamy Syndrome) caused by mutations of ARSB [17,18]. Accumulation of chondroitin sulfate is recognized as a significant factor in lung fibrosis of different etiologies [19–22], and treatment with recombinant ARSB ameliorated cardiac fibrosis in an animal model [23].

Many reports have considered how the pathophysiology of Covid-19 relates to interference with the normal balance between effects of Angiotensin (Ang) II and Ang1-7, due to binding of the SARS-CoV-2 spike protein with ACE2 (angiotensin converting enzyme receptor 2). SARS-CoV-2 spike protein binding with ACE2 can inhibit the ACE2-mediated production of Ang1-7 and lead to imbalance between AngII and Ang1-7/Mas receptor effects. AngII causes vasoconstriction initiated by interaction with the AT1 receptor, and decline in opposing vasodilation, due to reduced production of Ang1-7 when ACE2 is bound by the SARS-CoV-2 spike protein, may predispose to unopposed signaling events and significant pathophysiology [24–27]. The impact of inhibition of the renal angiotensin system (RAS) by angiotensin-converting enzyme (ACE) inhibitors or by angiotensin receptor blockers (ARBs) has been considered in relation to treatment and outcome of COVID-19 infection, and studies are ongoing [28–30].

Prior work showed marked increases in expression of CHST15 and CHST11 in rat vascular endothelial cells following exposure to Angiotensin (Ang) II [1]. These increases were largely, but incompletely, inhibited by treatment with an angiotensin receptor blocker, and consistent with possible activation by ACE2. The studies in this report support a sequence of activation of cellular transcriptional events which proceed from direct effects of the interaction between the spike protein receptor binding domain (SPRBD) and the ACE2 receptor in human airway epithelial cells (AEC). Recognition of the usurpation of normal signaling mechanisms by the SARS-CoV-2 spike protein-ACE2 interaction suggests the potential benefit of some targeted pharmacological interventions based on these pathways. Focus on the specific impact of the spike protein receptor binding domain (SPRBD) interaction with ACE2 on distinct signaling, mechanisms which increase CHST15 and CHST11 expression and inhibit ARSB expression has enabled identification of specific interventions which may have clinical benefit. In the experiments which follow, normal, primary human small airway cells were treated with SPRBD with and without Interferon-β (IFNβ), which augments the expression of ACE2 [31,32], and, thereby, amplifies the impact of the interaction between the SPRBD and ACE2. Elucidation of the roles of phospho-p38MAPK, phospho-Smad3, and Rb phosphorylation provides novel insights into how viral usurpation of endogenous signaling pathways can produce sustained pathophysiological consequences in human cells.

## Results

### Chondroitin sulfate, carbohydrate sulfotransferase (CHST)15, and N-acetylgalactosamine-4-sulfatase (Arylsulfatase B; ARSB) in post-mortem Covid-19 lung tissue and airway epithelial cells

In post-mortem lung tissue of patients with COVID-19, immunostaining showed decline in intensity and distribution of ARSB, in contrast to increases in total chondroitin sulfate in alveolar epithelial cells and in CHST15 in vascular-associated cells [1]. Previously published representative images from Covid-19 lungs and control tissue of total chondroitin sulfate (**Fig.1a,1b**), ARSB (**Fig.1c,1d**), and CHST15 (**Fig.1e,1f**) are reproduced [1]. Increased CHST15 was prominent in vascular smooth muscle cells of the Covid-19 lung tissue, compared to control tissue (**Fig.1g,1h**).

**Fig. 1.**
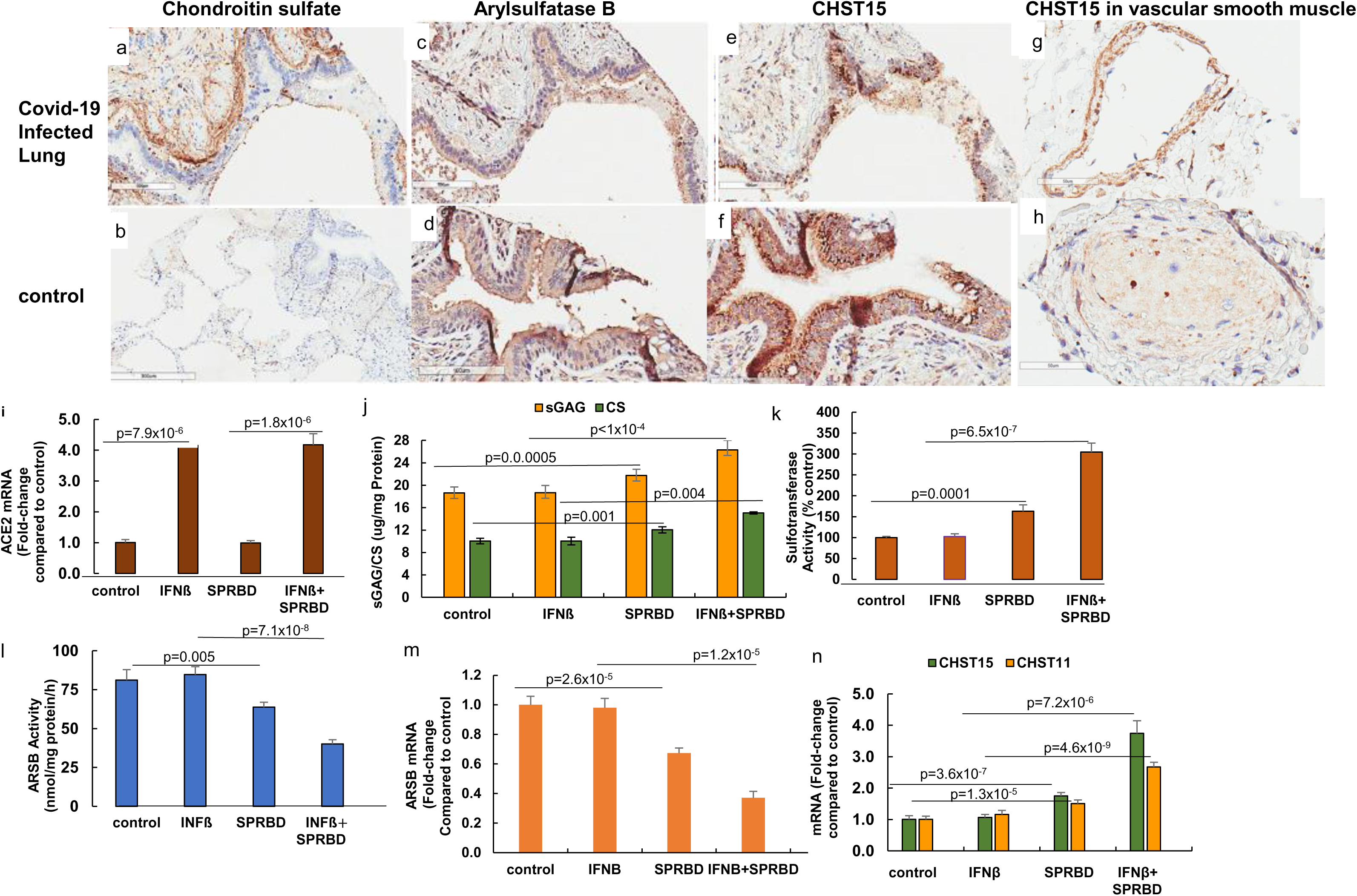
Chondroitin sulfate, Arylsulfatase B, and CHST15 in Covid-19 and normal control lung tissue and in human small airway epithelial cells. **a,b.** In lung tissue obtained post-mortem from patients who died due to SARS-CoV-2 infection, immunohistochemistry showed increased total chondroitin sulfate compared to normal control lung tissue. **c,d.** In contrast, intensity and distribution of ARSB were markedly reduced in the Covid-19 lung tissue. Marked decline is evident in the membrane immunostaining, compared to the normal control. **e,f.** Carbohydrate sulfotransferase (CHST) 15 which is the sulfotransferase that adds a 6-sulfate group to chondroitin 4-sulfate to form chondroitin-4,6-sulfate (chondroitin sulfate E), has regions of marked intensity of immunostaining in both the Covid-19 lung and the normal lung. **g,h.** In vascular smooth muscle tissue of the Covid-19 lung, the CHST15 immunostaining is much more intense and less diffuse than in the normal lung vascular tissue. **i.** Treatment of the cultured cells with Interferon (IFN)β amplifies the impact of SPRBD by increasing the expression of ACE2 more than four times the control level (p<0.1×10^−5^, n=6). **j.** Corresponding to the findings in the infected lung tissue, total chondroitin sulfate (CS) increased by ~2 ug/g protein (p=0.001, n=6) and total sulfated glycosaminoglycans increased by over 3 ug/g protein (sGAG) (p=0.0005, n=6) in the AEC following exposure to the SPRBD. Increases are more following combined treatment with SPRBD and IFNβ (>5 µg/g protein for CS, n=6; 7.7 µg/g protein for sGAG, n=6). **k.** Consistent with the observed increases in chondroitin sulfate in Covid-19 lung tissue and in the airway cells, sulfotransferase activity increased by 63% following SPRBD and by over 200% following the combination of SPRBD and IFNβ (n=6, n=6) **l.** Arylsulfatase B (ARSB) activity declined in the AEC following exposure to the SPRBD (p=0.005, n=6), and declined over 50% by the combined exposure to SPRBD and IFNβ. **m.** The mRNA expression of ARSB also declined (p=2.6×10^−5^; n=6) following SPRBD, and declined further following exposure to the combination of SPRBD and IFNβ. **n.** Expression of both CHST15 and of CHST11, which transfers 4-sulfate groups to N-acetylgalactosamine residues to create chondroitin 4-sulfate, is significantly upregulated following exposure to the SPRBD and the combination of SPRBD and IFNβ. All of the p-values were determined using unpaired t-test, two-tailed, with unequal variance, and error bars represent one standard deviation. [ACE2=angiotensin converting enzyme 2 receptor; AEC=normal, human small airway epithelial cells; ARSB=arylsulfatase B=N-acetylgalactosamine-4-sulfatase; SPRBD=SARS CoV-2 spike protein receptor binding domain]

Normal, primary human, small airway epithelial cells (AEC) were treated with the SARS-CoV-19 spike protein receptor binding domain (SPRBD; 2.5 µg/ml x 2h), following exposure to Interferon-β (IFNβ; 10 ng/ml x 24h), which was used to increase the expression of angiotensin converting enzyme 2 (ACE2) receptor, and thereby amplify the effect of SPRBD and better simulate the impact of viral infection. The expression of ACE2 increased to more than four times the baseline following IFNβ or the combination of IFNβ and SPRBD, and SPRBD alone did not have any significant impact on ACE2 (**Fig.1i**). Consistent with the observed findings from immunohistochemistry, total chondroitin sulfate and total sulfated glycosaminoglycans (GAGs) increased significantly following exposure to SPRBD and to the combination of SPRBD and IFNβ. Almost two thirds of the increase in the sulfated GAGs was due to the increase by over 5 ug/mg protein of chondroitin sulfate (**Fig.1j**). Sulfotransferase activity increased by over 200% following exposure to the SPRBD and IFNβ (**Fig.1k**). ARSB activity (**Fig.1l**) and mRNA expression (**Fig.1m**) declined following exposure to the SPRBD. The mRNA expression of carbohydrate sulfotransferases CHST15 and CHST11 increased to 3.8 and 2.7 times the control values following exposure to SPRBD and IFNβ (p<0.001, p<0.001; n=6) (**Fig.1n**).

### Inhibition of phospho-Thr180/Tyr182-p38-MAPK or of phospho-S423/S425-SMAD3 blocks increases in CHST15 and CHST11

The cultured AEC were treated with inhibitors of cell signaling, including SB203580, an inhibitor of phospho-(Thr180/Tyr182)-p38 MAPK, and NSC23766, a Rho/Rac-1 GTPase inhibitor (**Fig.2a**). Marked declines in expression of CHST15 and CHST11 followed exposure to the phospho-(Thr180/Tyr182)-p38 inhibitor, but NSC23766 had no effect. Increases in CHST15 and CHST11 expression were blocked by SIS3 (specific inhibitor of SMAD3) (**Fig.2b**). Effectiveness of ACE2 silencing was confirmed by QPCR (**Fig.2c**), and the impact of silencing ACE2 tested to confirm that ACE2 was required for the observed effect of the SPRBD. ACE2 siRNA blocked the SPRBD-induced increase in phospho-(Thr180/Tyr182)-p38 (**Fig.2d**), demonstrating dependence on ACE2 for the observed effects. As expected, the increase in phospho-(S423/S425)-SMAD3 following exposure to SPRBD was inhibited by SIS3 (**Fig.2e**), and was also partially inhibited by SB203580 (**Fig.2e**), indicating participation by phospho-p38 in the SPRBD-induced increase in phospho-SMAD3. Phospho-(Thr180/Tyr182)-p38 MAPK was significantly increased following exposure of the cells to SPRBD (**Fig.2f**), but SIS3 had no impact on the increase. Increases in promoter activation of CHST15 (**Fig.2g**) and CHST11 (**Fig.2h**) were abrogated by treatment with either SIS3 or SB203580 following exposure to SPRBD.

**Fig. 2.**
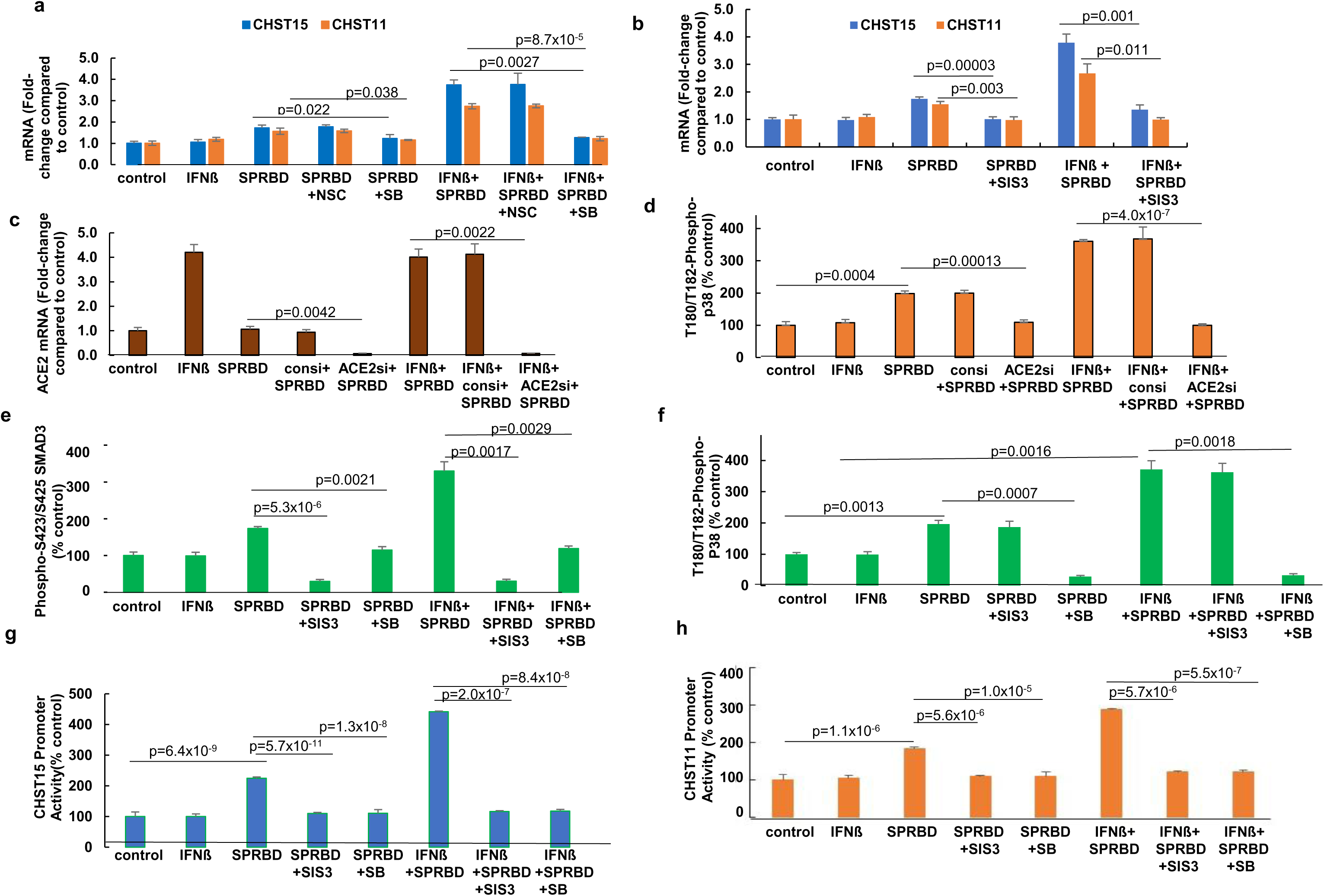
SMAD3 and p38-MAPK inhibitors abrogate the effects of SPRBD. **a.** The SPRBD- and SPRBD+IFNβ-induced increases in the mRNA expression of CHST15 and CHST11 were significantly reduced following exposure to SB20850, the p38-MAPK inhibitor. In contrast, NSC23766, a Rho/Rac-1 GTPase inhibitor, had no impact on their expression. **b**. SIS3, an inhibitor of phospho-Smad3, inhibited the SPRBD- and SPRBD+IFNβ-induced increases in expression of CHST15 and CHST11. **c.** Expression of ACE2 was inhibited by ACE2 specific siRNA following exposure to SPRBD alone or with IFNβ. **d.** Following silencing of ACE2 by siRNA, the SPRBD- and SPRBD+IFNβ-induced increases in phospho-T180/T182-p38 MAPK were inhibited. **e**. The SPRBD- and SPRBD+IFNβ-induced increases in phospho-S423/S425 SMAD3 were completely inhibited by SIS3 and, to a lesser extent, by SB20850, the p38 MAPK inhibitor. **f.** The SPRBD and SPRBD+IFNβ-induced increases in phospho-Thr180/Tyr182-p38 were unaffected by exposure to SIS3 and were inhibited by SB20850. **g,h.** Promoter activity of CHST15 and CHST11 was enhanced by exposure to SPRBD and to a greater extent by the combination of IFNβ and SPRBD. Both SIS3 and SB20850 inhibited promoter activation. [SIS3=specific inhibitor of SMAD3]. All p-values were determined by unpaired t-test, two-tailed, with unequal variance, and with at least three independent experiments. Error bars represent one standard deviation.

### Phospho-p38 MAPK mediates decline in ARSB expression through effects on Rb phosphorylation and E2F1

In contrast to the observed increases in CHST11 and CHST15, the mRNA expression and activity of ARSB declined significantly following exposure to the SPRBD (**Fig.1l,1m**). ARSB expression increased following treatment by SB203580 (**Fig.3a**). Neither NSC23766 (**Fig.3a**) nor SIS3 (**Fig.3b**) reversed the effect of SPRBD. ARSB promoter activation was reduced by SPRBD exposure, and treatment with SB203580 reversed the decline in ARSB (**Fig.3c**).

**Fig. 3.**
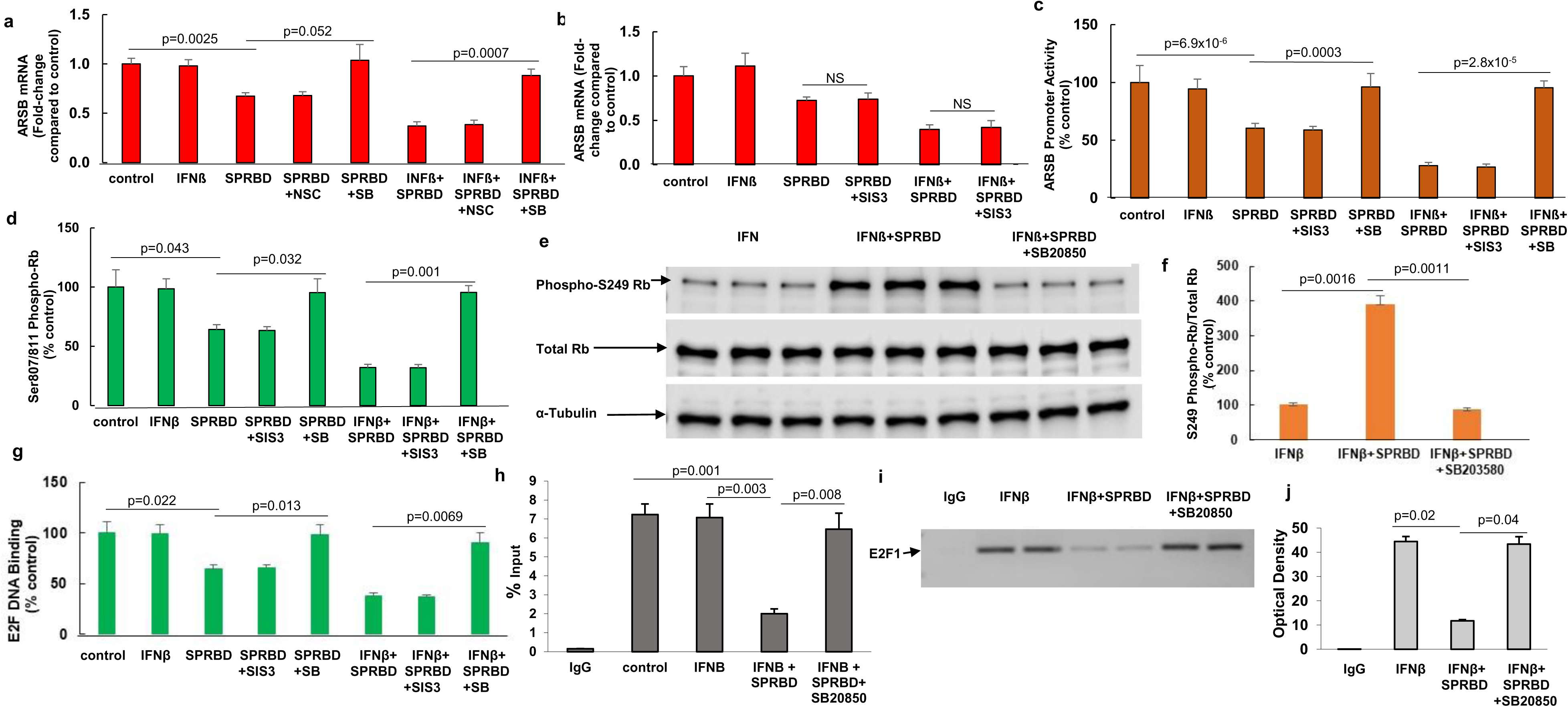
Exposure to spike protein receptor binding domain inhibits ARSB expression by activation of phospho-p38 MAPK and phospho-(S249/T252)-RB-E2F1 interaction. **a.** ARSB mRNA expression was unaffected by NSC23766, but following exposure to the p38-MAPK inhibitor SB20850, mRNA expression was restored to baseline control values. **b.** In contrast, SIS3 had no impact on the SPRBD- or SPRBD+IFNβ-induced decline in ARSB expression. **c.** ARSB promoter activation was reduced by SPRBD and by SPRBD+IFNβ. These declines were reversed by exposure to SB20850, but not by SIS3. **d.** Following treatment with SPRBD or SPRBD+IFNβ, C-terminal phospho-(Ser807/811)-Rb declined as shown by ELISA. These declines were reversed by SB203580, but not by SIS3. **e.** In contrast to the decline in C-terminal Rb phosphorylation, exposure to SPRBD+IFNβ increased N-terminus phospho-(S249)-Rb, as detected by Western blot. This increase was inhibited by SB20850, and total Rb was unchanged. **f.** Densitometry confirms the impression of Western blot and shows the ratio of phospho-S249-Rb to total Rb following SPRBD+IFNβ has increased to 3.89 times the baseline. **g**. Following exposure to SPRBD and SPRBD+IFNβ, E2F-DNA binding declined significantly, and SB20850 reversed the declines. **h.** %DNA input declined following SPRBD+IFNβ and increased following inhibition of p38-MAPK by SB20850. **i.** Chromatin immunoprecipitation (ChIP) shows decline in E2F1 binding to the ARSB promoter following exposure to SPRBD+IFNβ; decline was reversed by SB203580. Agarose gel demonstrates no effect of the IgG negative control on E2F1 binding to the ARSB promoter at baseline, IFNβ control, reduced binding following SPRBD+IFNβ, and reversal of the decline following treatment with SB20850. **j**. Measurements of optical density confirm the impression from the gel. P-values were determined by unpaired t-tests, two-tailed with unequal variance, with n of at least 3 independent experiments. Error bars show one standard deviation.

Since phospho-p38-MAPK was reported to activate N-terminal Rb (retinoblastoma protein) phosphorylation [33,34] and thereby inhibit E2F1-DNA binding and the ARSB promoter has multiple E2F1 binding sites [35], the impact of the p38-MAPK inhibitor SB203580 on Rb phosphorylation following exposure to SPRBD was addressed in the AEC. SB203580 reversed the SPRBD-induced decline in C-terminal phospho-(Ser807/811)-Rb (**Fig.3d**). In contrast, SB203580 reduced the SPRBD-induced increase in N-terminal Rb phosphorylation (phospho - Ser249/Thr252), as shown by Western blot (**Fig.3e**) and corresponding values of phospho- (S249/Thr252)-Rb to total Rb (**Fig.3f**). The phospho-(S249/T252)-Rb to total Rb ratio increased by exposure to SPRBD and declined following SB203580, reflecting the dependence of N-terminal Rb phosphorylation on phospho-p38. Since N-terminal Rb-phosphorylation, in contrast to C-terminal Rb phosphorylation, activates Rb binding with E2F1 [33,34] and reduces availability of E2F for DNA binding, the impact of SPRBD on E2F-DNA binding was assessed. E2F transcription factor-DNA binding assay showed decline following SPRBD-IFNβ exposure and recovery following SB203580 (**Fig.3g**). Chromatin immunoprecipitation (ChIP) assay was performed to assess specific E2F1-binding to the ARSB promoter. Percent DNA input declined following SPRBD-IFNβ exposure, but increased following SB203580 (**Fig.3h**). Agarose gel shows that E2F1-DNA binding to the ARSB promoter was reduced by SPRBD-IFNβ exposure, but was reversed by SB203580 (**Fig.3i**), as confirmed by densitometry (**Fig.3j**). These findings implicate a complex signaling mechanism whereby phospho-p38 MAPK phosphorylates N-terminal Rb, thereby enhancing Rb-E2F1 binding and negatively regulating the ARSB promoter and suppressing ARSB expression.

### Effects of Desloratadine, Monensin, and Dexamethasone on expression of CHST15, CHST11, and ARSB

Multiple treatments have been investigated in relation to impact on mortality and morbidity from Covid-19 infection. Both H1 and H2 antihistamine receptor antagonists have been considered, including the H1 antihistamine loratadine and its metabolite desloratadine [36–38]. Treatment of SPRBD-exposed AEC by desloratadine reduced the SPRBD-induced increase of phospho-Thr180/Tyr182-p38-MAPK (**Fig.4a**) and the mRNA expression of CHST15 and CHST11 (**Fig.4b**). Desloratadine also countered the SPRBD-induced decline in ARSB expression (**Fig.4c**) and activity (**Fig.4d**).

**Fig. 4.**
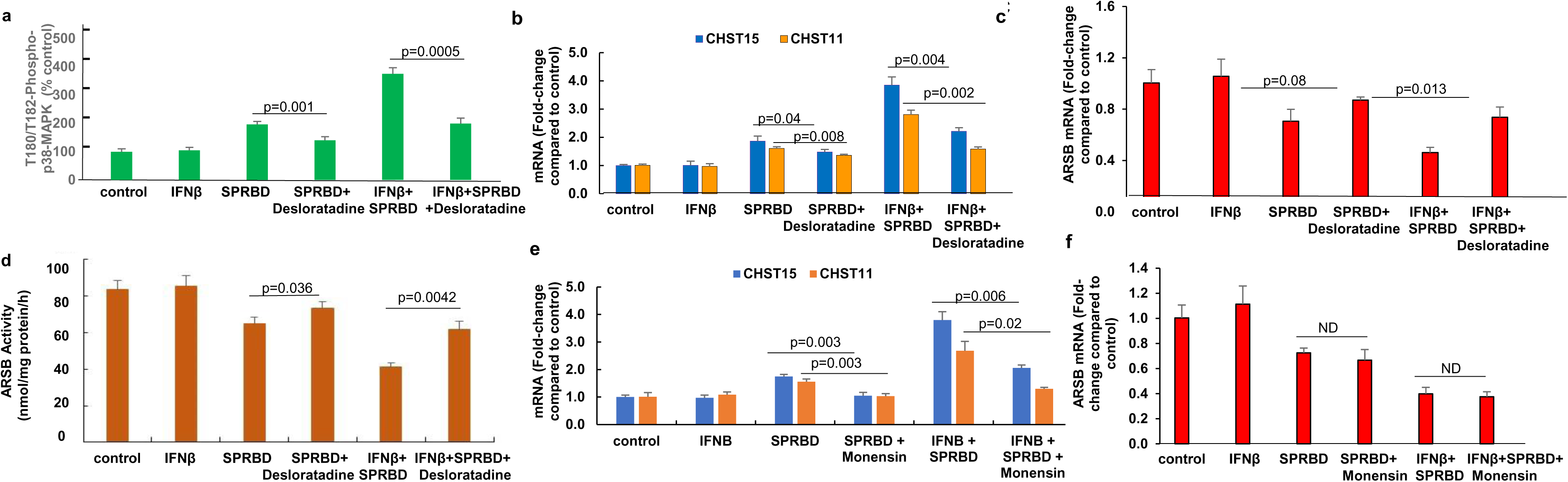
Effects of selected agents on expression of CHST15, CHST11, and ARSB. **a.** The antihistamine desloratadine reduced by 62% the SPRBD-induced increase in T180/T182 phospho-p38-MAPK in the AEC. **B.** Consistent with the decline in phospho-p38, desloratadine reduced the SPRBD-induced and SPRBD with IFNβ-induced increases in CHST15 (p=0.045, p=0.0041 with IFNβ) and CHST11 expression (p=0.008, p=0.0017 with IFNβ, unpaired t-tests, two-tailed, unequal variance, n=3). **C, D.** Consistent with the observed decline in ARSB, desloratadine partially reversed the SPRBD-induced decline in ARSB activity (from 64.7 to 73.4 nmol/mg protein/h and from 41.0 to 61.8 nmol/mg protein/h with IFNβ; n=3) and expression (0.71 to 0.87 and 0.46 to 0.74 with IFNβ, fold-change compared to control; n=3). **E.** The polyether antibiotic monensin, which modifies dermatan sulfate biosynthesis and processing [39–41], reduced the SPRBD-induced increases in CHST15 mRNA (p=0.003, p=0.006 with IFNβ) and CHST11 (p=0.003, p=0.002 with IFNβ; fold-change compared to control, unpaired t-tests, two-tailed, unequal variance). **F**. Monensin had no significant impact on the SPRBD-induced decline in ARSB.

The polyether antibiotic monensin, which has been shown to have effects on sulfated glycosaminoglycan biosynthesis [39–41], reduced the SPRBD-induced increases in mRNA expression of CHST15 and CHST11 (**Fig.4e**), but had no effect on the SPRBD-induced decline in ARSB (**Fig.4f**). Dexamethasone, which has been widely used clinically, did not reverse effects of SPRBD on CHST15, CHST11, or ARSB expression.

## Discussion

In this report of *in vitro* effects following exposure of cultured normal, human small airway epithelial cells to the SARS-CoV-2 spike protein binding domain, data demonstrate increased expression of chondroitin sulfotransferases CHST15 and CHST11, increased sulfotransferase activity and increased content of chondroitin sulfate, and decline in ARSB expression and activity. Reduced expression of ARSB leads to the accumulation of chondroitin sulfate, since hydrolysis of the 4-sulfate group is required for chondroitin 4-sulfate degradation [42–45]. Both the increases in CHST15 and CHST11 and the decline in ARSB are mediated by phospho-p38 MAPK, and SMAD3 is required for CHST15 and CHST11 expression, whereas the expression of ARSB is regulated by N-terminus Rb phosphorylation and E2F1-Rb interaction, as presented in **Fig.5**. Detailed studies of N-terminal Rb phosphorylation [33,34] present a complex mechanism by which Rb-E2F binding is increased, rather than decreased by C-terminal Rb-phosphorylation. Application of this mechanism of Rb activation by N-terminal phosphorylation, which overrides Rb inhibition by C-terminal phosphorylations, provides a novel, coherent approach to how ARSB expression may be down-regulated following stimulation of phospho-p38 by SPRBD-ACE2 in the AEC.

**Fig. 5.**
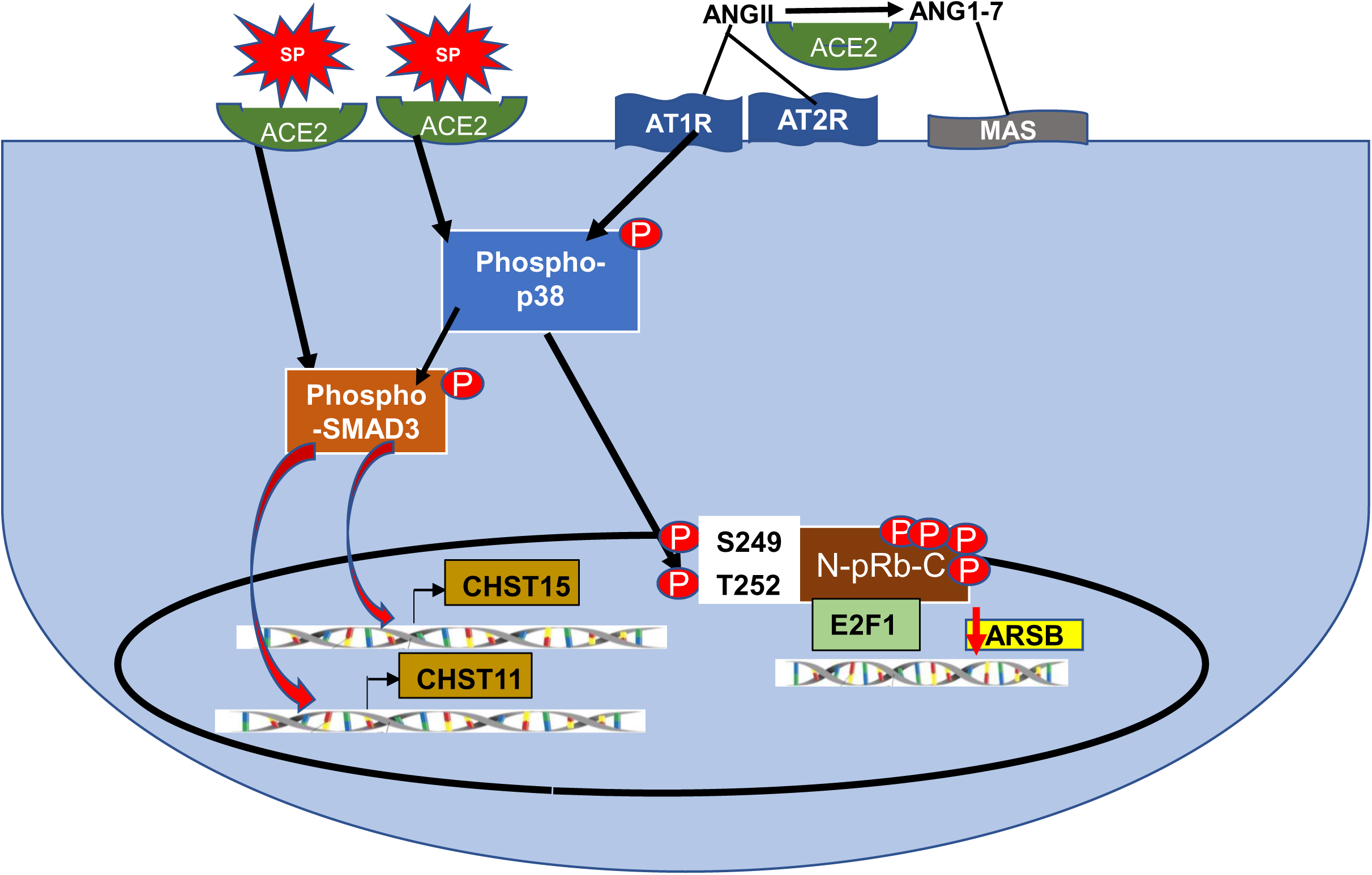
SARS-CoV-2 spike protein binding with ACE2 initiates transcriptional events which modify chondroitin sulfation through activation of phospho-p38 MAPK. Schematic of SARS-CoV-2 spike protein binding domain (SPRBD) peptide interaction and initiation of transcriptional events which modify chondroitin sulfation through activation of phospho-p38 MAPK. Both p38 MAPK and phospho-Smad3 lead to increased expression of CHST15 and CHST11. AngII interaction with AT1 is also expected to increase phospho-p38. Phospho-p38 leads to N-terminal phosphorylation and activation of Rb with enhanced binding of E2F1, which reduces ARSB promoter activation due to reduced binding of E2F1. [ACE2=angiotensin converting enzyme 2; AngII=angiotensin II; ARSB=arylsulfatase B; AT1R=angiotensin II receptor type 1; AT2R=angiotensin II receptor type 2; CHST=carbohydrate sulfotransferase; pRb=retinoblastoma protein; SMAD=Suppressor of Mothers against Decapentaplegic; SPRBD=SARS-CoV-2 spike protein receptor binding domain]

Study results indicate that pathways leading to decline in ARSB and increases in expression of CHST15 and CHST11 following exposure to the SPRBD require activation of phospho-p38 MAPK. P38-MAPK has been implicated in critical interactions between exogenous exposures, intracellular signaling, and transcriptional events in a wide range of experiments [46–49]. Crosstalk between the p38 and Smad pathways is involved in fibrogenic signaling following AngII and downstream of TGF-β [50–52]. The current experiments provide novel insight into how phospho-p38 participates in the regulation of the biosynthesis and maintenance of 4-sulfated chondroitins.

Other work has shown that when ARSB is inhibited and 4-sulfation sustained, SHP2 binding to chondroitin 4-sulfate is increased, leading to reduced SHP2 activity and sustained phosphorylation of ERK1,2, JNK, and p38 [14,53–56]. The impact of phospho-38-MAPK on N-terminal Rb phosphorylation, Rb-E2F1 binding, and suppression of ARSB promoter activation, may be part of a loop, in which p38 phosphorylation is sustained, due to enhanced chondroitin 4-sulfate-SHP2 binding which follows decline in ARSB and increased chondroitin 4-sulfation.

Increased understanding of how chondroitin sulfate is involved in the pathogenesis of Covid-19 may help in the development of preventive and therapeutic strategies. The potential benefit of the H1 histamine receptor blocker Desloratadine is suggested by the measured decline in phospho-p38 and by inhibition of the SPRBD effects on expression of CHST15, CHST11 and ARSB, as well as by pharmacological studies that identified desloratadine/loratadine as a mechanism-based target [36–38]. Several clinical studies of H2 blockers on the response in Covid-19 have been reported and additional studies of antihistamines are ongoing [57–59].

Analysis of intensity and distribution of total chondroitin sulfate, CHST15 and ARSB immunohistochemistry in sections from SARS-CoV-2-infected lungs demonstrated marked increase in chondroitin sulfate and prominence of vascular-associated CHST15, in contrast to decline in ARSB [1]. These findings were similar to those observed in diffuse alveolar damage from other causes and suggest that accumulation of chondroitin sulfate might be a significant component in refractory lung disease and pulmonary fibrosis. Previously, decline in ARSB was implicated in the response to hypoxia in human bronchial epithelial cells [60] and in failure of patients with moderate COPD to respond to oxygen therapy [61]. These findings provide additional evidence that decline in ARSB contributes to refractory clinical response in SARS-CoV-2 infection.

By use of the specific spike protein receptor binding domain which binds with ACE2, the experiments in this report have focused on how disruption of normal ACE2 function by the SARS-CoV-2-ACE2 interaction affects signaling in normal human airway cells. In this model, viral uptake does not occur, and other spike protein mediated interactions, such as with TMPRSS2, are not anticipated. A specific p38 isoform, such as p38α, may predominate in the reactions presented in this report, and future work will help to clarify which p38 isoform(s) is most involved. Also, the precise mechanism by which the SPRBD-ACE2 interaction leads to activation of phospho-p38-MAPK is not yet clarified. Interception of the activation of p38-MAPK emerges as a therapeutic goal, and H1-receptor blockers may directly inhibit phospho-p38 activation and, thereby, prevent the effects of increased expression of CHST15 and CHST11, reduced expression of ARSB, and accumulation of excess chondroitin sulfate following SARS-CoV-2 infection. Also, atypical antibiotics, such an monensin, which directly affect chondroitin 4-sulfate and dermatan sulfate synthesis, may provide new targets to disrupt virus-initiated signaling. Increased attention to the impact of chondroitin sulfates and ARSB in Covid-19 pathobiology and the role of phospho-p38 activation in the mechanisms of their expression may yield new tools and new insights which will reduce the morbidity and mortality of Covid-19.

## Author Contributions

JKT and SB designed the experiments, SB performed the experiments, and wrote the manuscript.

## Data Availability

All data produced in the present study are available upon reasonable request to the authors.

